# Microbiome profiles are associated with cognitive functioning in 45-month-old children

**DOI:** 10.1101/2021.06.26.21259573

**Authors:** Fabian Streit, Emese Prandovszky, Tabea Send, Lea Zillich, Josef Frank, Sarven Sabunciyan, Jerome Foo, Lea Sirignano, Bettina Lange, Svenja Bardtke, Glen Hatfield, Stephanie H Witt, Maria Gilles, Marcella Rietschel, Michael Deuschle, Robert Yolken

## Abstract

Prenatal, perinatal, and postnatal factors have been shown to shape neurobiological functioning and alter the risk for mental disorders later in life. The gut microbiome is established early in life, and interacts with the brain via the brain-immune-gut axis. However, little is known about how the microbiome relates to early-life cognitive functioning in children. The present study, where the fecal microbiome of 380 children was characterized using 16s rDNA and metagenomic sequencing aimed to investigate the association between the microbiota and cognitive functioning of children at the age of 45 months measured with the Wechsler Preschool and Primary Scale of Intelligence (WPPSI-III).

Overall the microbiome profile showed a significant association with cognitive functioning. A strong correlation was found between cognitive functioning and the relative abundance of an unidentified genus of the family *Enterobacteriaceae*. Follow-up mediation analyses revealed significant mediation effects of the level of this genus on the association of maternal smoking during pregnancy and current cigarette smoking with cognitive function. Metagenomic sequencing of a subset of these samples indicated that the identified genus was most closely related to *Enterobacter asburiae*. Analysis of metabolic potential showed a nominally significant association of cognitive functioning with the microbial norspermidine biosynthesis pathway.

Our results indicate that alteration of the gut microflora is associated with cognitive functioning in childhood. Furthermore, they suggest that the altered microflora might interact with other environmental factors such as maternal cigarette smoking. Interventions directed at altering the microbiome should be explored in terms of improving cognitive functioning in young children.

## Introduction

The microbiome, a collection of genes of the multitude of microorganisms living in and on the human body, has been shown to play a crucial role in somatic health (1,2). Recently, its role in the etiology of mental disorders has been documented by a large body of research. The gut microflora has been demonstrated to interact with the brain (gut-brain axis), via neural (vagus nerve), endocrine, immune and humoral (cytokines, short-chain and long-chain fatty acids) reciprocal communication routes (3-5). Moreover, it has been implicated in mental health and has been linked to disorders such as schizophrenia, bipolar disorder, attention deficit hyperactivity disorder, and autism spectrum disorders (6-8). Modern sequencing approaches make it possible to investigate these dense microbial populations, such as those prevalent in the human gut (1). It has been found that the microbial composition varies between both individuals (9,10) and different body sites (1). The human microbiome undergoes major changes in the first 1–3 years of life (11,12) but then remains relatively stable on an individual level (9,13,14). However, it has yet to be clarified whether bacterial colonization of the human body starts in utero or during birth (15). During the birth process, infants are exposed to a variety of maternal and environmental microbiota. The microbiome can be influenced by various factors including the mode of delivery (16), breast vs. formula feeding (14,17), and nutrition in general as well as the use of medication, especially antibiotics (18-20).

While the first few years of life are crucial for the establishment of the microbiome, this period is also a critical phase for the development of neurobiological functioning and is likely to represent a critical phase in the etiology of neurodevelopmental psychiatric disorders (21). Cognitive functioning in children highly reflects healthy neurodevelopment and is related to a broad range of mental health outcomes (22,23). Several animal studies have demonstrated an interaction between the gut microbiome and cognitive functioning (24-26). Studies have also linked alterations in the microbiome composition to cognitive functioning in humans (27). It was recently reported that the microbiome of children measured at one year of age correlated with cognitive functioning and brain volume at age one and two (28) an association which has been confirmed in other studies (29-31). However, only a few studies have discussed the relationship between the microbiome and cognitive functioning in older infants and young children, where mental health and cognitive outcomes are already more advanced and easier to evaluate. The pre-school time period between three and five years of age is of particular interest due to its importance in subsequent learning and cognitive development (32).

In young children many factors can contribute to changes in the microbiome and might subsequently have an impact on cognition later in life. For example, exposure to antibiotics —a major modulator of the microflora— in the first six months of life has been linked to lower cognitive abilities at 11 years of age (33). Additional factors that have been shown to affect the microflora during infancy including mode of delivery (Caesarean section vs. vaginal birth), type of feeding (breastfeeding vs. formula), gestational age, diet, and socioeconomic status (34-36). Maternal smoking is another major neurodevelopmental risk factor during pregnancy, and has previously been linked to reduced birth weight in this cohort (37) and in others (38). However, the effect of these factors on the microbiome and cognitive functioning in the pre-school aged children has not been extensively studied (39).

In the present study, we aimed to I) investigate the link between cognitive functioning and the intestinal microbiome at the age of 45 months, II) identify particular bacteria species and microbial metabolic pathways that are associated with cognitive performance, and III) test whether the most strongly associated covariate (smoking during pregnancy) influences theses associations.

## Methods

### Study population

Study participants were part of an ongoing longitudinal study on pre-, peri-, and postnatal stress and offspring development and health (Pre-, Peri-, and Postnatal Stress: Epigenetic Impact on Depression; POSEIDON) (40-42). In total, 410 pregnant women were enrolled in the study around four to eight weeks prior to delivery. Inclusion criteria for the mothers were: age 16–45 years, German-speaking, and being the child’s primary caregiver. Exclusion criteria were evidence of infection with hepatitis B, or hepatitis C or human immunodeficiency virus, current inpatient treatment due to a diagnosis of a psychiatric disorder, a lifetime diagnosis of schizophrenia or psychotic disorder, or a substance dependency other than nicotine during pregnancy. For children, the exclusion criteria were birth before a gestational age of 30 weeks, birth weight lower than 1,500 grams, or multiple births, as well as any congenital disease, malformation, or chromosomal abnormality. The study was registered in the German Clinical Trials Register (DRKS00006338), approved by the Ethics Committee of the Medical Faculty Mannheim of the University of Heidelberg and conducted in accordance with the Declaration of Helsinki. All families provided written informed consent.

To date, four waves of assessments have been carried out: T1 (during the third trimester of pregnancy), T2 (between day 1–3 after birth), T3 (at the age of 6 months), and T4 (at the age of 45 months). At T4, of the original 410 families, 302 remained, and the dropouts were replaced with 101 newly recruited families. For the newly recruited families, pre-, peri-, and early postnatal factors were coded based on parental reports or medical records. Details of the assessment procedure have been described previously (42-44).

Assessment included the following variables used in the present study: smoking during pregnancy assessed via self-report, Caesarean section (C-section) or natural birth, breastfeeding, age of the mother as well as sex, birth weight, and gestational age of the child. Measures at T4 used in the present study included the child’s age and BMI, use of antibiotic medication during the previous 6 months, current maternal smoking, multilingual upbringing, and the years of maternal education as a marker of socioeconomic status (45). Perceived stress ratings of the mothers using Perceived Stress Scale (PSS) (Cohen, Kamarck, & Mermelstein, 1983) were available during pregnancy for the original sample only, and at T4 for the original and the newly recruited sample.

Cognitive functioning was assessed using the Wechsler Preschool and Primary Scale of Intelligence - Third Edition (WPPSI-III) (46) at T4, the same time point at which stool sampling occurred. Five subtests of the WPPSI-III were conducted: the core subtests *Receptive Vocabulary, Information, Block Design*, and *Object Assembly*, and the supplementary subtest *Picture Naming*. For all subtests, the raw scores were converted to standardized scores based on the child’s exact age. Full-scale IQ (FIQ) score was calculated from the four core subtests, verbal IQ (VIQ) from the *Receptive Vocabulary* and *Information* subtests, performance IQ (PIQ) from the *Block Design* and *Object Assembly* subtests, and the general language composite (GLC) from the *Receptive Vocabulary* and *Picture Naming* subtests. Subjects were included in the analysis if values were available for at least one of the WPPSI-III scales, resulting in the exclusion of 21 subjects. Prior to the statistical analyses, the test documentation was reevaluated as an additional quality control step. Data points for each subtest were excluded if, for example, an apparent lack of motivation or lack of understanding the instructions was evident. This reevaluation resulted in the exclusion of additional 14 subjects. Five more samples were excluded due to irregularities in shipping or handling (e.g. delayed shipping, extended storage at home before shipping).

### Microbiome analysis

#### Sampling and storage

Stool samples were collected by the parents at the age of 45 months (T4). The samples were collected using a standard stool sample kit (Sarstedt, Nuembrecht, Germany) on the days prior to or the day of the T4 appointment and temporarily stored in a home refrigerator until being delivered by the parents at ambient temperature to the lab on the day of the appointment. In circumstances when this was not achievable, stool samples were sent to the laboratory at ambient temperature by mail service, which usually took 1-2 days. The effect of the shipment on the microflora was analyzed and taken into account as noted below. Upon arrival, stool samples were frozen at -80°C and stored in the Biobank of Psychiatric Diseases Mannheim (BioPsy) (47) until further processing.

#### DNA extraction

Fecal samples were thawed and an aliquot of 100 mg was used to extract genomic DNA using the ZymoBIOMICS^™^ 96 MagBead DNA Kit according to the manufacturer’s protocol. The protocol included a bead-beating step to enable complete homogenization/disruption of the microbial cell walls. ZymoBIOMICS^™^ Microbial Community Standard (Zymo Research Europe GmbH) was included in the DNA extraction process to ensure an accurate representation of microbial communities in samples (48) and to assess potential environmental contaminants. DNA quantity and quality were determined by Thermo Scientific NanoDrop^™^ 1000 Spectrophotometer and normalized for further analysis to 5 ng/µL.

#### Characterization of the Microbiome

##### Library preparation

The extracted DNA was amplified using primers directed at conserved bacterial 16S ribosomal sequences in the v3–v4 region, and 380 individually indexed (Nextera XT, Illumina, San Diego, CA) libraries were generated following the Illumina 16S metagenomic library preparation protocol. Each library was quantified using Qubit (Life Technologies) and the concentration was normalized to 4 □nM. A 5□μl aliquot of each library was pooled and denatured using 0.2 □N NaOH and subsequently diluted to 10 □pM using HT1 (hybridization buffer) supplied by Illumina (San Diego, CA). Prior to loading, the library was spiked with a 10% 20 □pM PhiX control. Sequencing was performed on the MiSeq Illumina sequencing platform using a 2 × 300 PE sequencing reagent kit (Cat # MS-102-3003 Illumina, San Diego, CA).

##### Sequence Analysis

Raw paired-end reads were first de-multiplexed using 5′ index information, and adapter sequences were trimmed off prior access. Sequence reads were analyzed separately using the QIIME 2 (v 2018.11) platform (49). In addition to filtering out any remaining PhiX contaminants and chimeric sequences, the DADA-q2 plugin (50) was utilized to remove low-quality reads. All unique sequence reads were collected into a feature table, which was filtered to remove low-abundance features using the feature*-*table q2 plugin (51). Next, multiple sequence alignment was performed by employing the MAFFT alignment plugin (52) followed by computing a phylogenetic tree using the FastTree program (53). Sampling depth was set to 2263 sequence reads. Phylogenetic diversity analyses utilized this phylogenetic tree in order to calculate the following alpha diversity matrices: number of observed species, Shannon’s and Faith’s phylogenetic diversity, and Pileou’s Evenness. To explore the bacterial composition of the samples, taxonomy was assigned to sequences using a pre-trained naive Bayes classifier (54) and q2-feature-classifier plugin (https://github.com/qiime2/q2-feature-classifier). This classifier was trained on Greengenes v13_8 99% OTUs, where the sequences have been trimmed to only include v3–v4 regions of the 16S rRNA gene bound by Illumina primer pairs. Analyses of taxa focused on the level of genera since that is the taxonomic level that can be determined with the highest degree of certainty with this method (55). Prior to analysis, the number of counts for each genus were log-transformed using natural logarithm after adding 1, to allow for the analysis of samples with 0 counts.

##### Metagenomic Shotgun Sequencing

A subset of 39 samples and 1 blank were selected for metagenomic sequencing. The selection was based on a broad distribution of WPSSI-III full scale IQ score and *Enterobacteriaceae* genus abundance, including samples with both low and high values. After exclusion for invalid WPSSI values or irregularities in sample shipping or handling, data from 33 subjects was used for statistical analysis. Aliquots of 1ng of extracted fecal DNA from the selected samples were used to generate paired-end metagenomic libraries using Illumina Nextera XT Low Input DNA kit following the manufacturer’s instructions (<u>Nextera XT DNA Library Prep Kit Reference Guide (15031942)</u>). The concentration of each library was measured by using Quibit and normalized to 4nM. Libraries were then sequenced using the Illumina HiSeq 4000, using the sequencing core facility at University of Maryland. Sequence reads with homology to human sequences were removed according to the selected downstream pipeline. The remaining sequence reads were then mapped to microbial databases employing Centrifuge (v1.0.3) (56) as previously described. Centrifuge uses a novel indexing scheme based on the Burrows-Wheeler transform (BWT) and the Ferragina-Manzini (FM) index optimized specifically for the metagenomic classification problem. Centrifuge output was visualized using the R-based application Pavian (https://ccb.jhu.edu/software/pavian/) (57). The metabolic potential of the microbiome was characterized using the program humann2 (v1.7) as described elsewhere (58). For association analysis with cognition, we selected a set of pathways, I.) focusing on the pathway abundances at community level, II.) filtering hits present in the blank sample, or present in more than 37 and less than 5 samples and III.) with a variance at least 3 times higher than the average abundance. This resulted in a list of 122 pathways (see Supplementary Table S6).

#### Statistical Analyses

Restricted maximum likelihood (REML) models were computed to assess the association of the overall microbiome profile with the cognitive measures as implemented in the software package omic-data-based complex trait analysis (OSCA v.0.45) (59). This approach applies REML to assess the overall association of the microbiome profile with the tested variables, expressed as the share of explained variance (59). Corresponding 95% confidence intervals (CI) were calculated with bootstrapping as implemented in FIESTA v1.0 (61). In a second step, potentially confounding covariates for those analyses were identified in separate REML model for each confounding variable: maternal age, sex of the child, infant feeding (formula vs. breast), gestational age, birth weight, antibiotic exposure during the 6 months prior to T4 (no vs. yes), type of birth (vaginal delivery vs. C-section), maternal cigarette smoking during pregnancy (none vs. any), current maternal cigarette smoking at T4 (none vs. any), child age at T4, child BMI at T4, multilingual upbringing at T4, level of maternal education (in years of education), and shipment vs. delivery by parents. Subsequently, the cognition REML models were rerun, adding confounding variables which showed a significant association with the microbiome profile as covariates.

Further analyses were conducted with the R open-source environment for statistical computation and graphics (v3.5.1) (R Development Core Team, 2013). Regression analyses were used to explore bivariate associations between the WPPSI-III full-scale IQ score and the following microbiome measures: alpha diversity measures, relative abundance of specific genera, and predicted metabolic pathways. Standardized betas (*ß)* and their 95% confidence intervals (CI) are given. The resulting *p* values were corrected for multiple comparisons (indicated as *q*-values) using Bonferroni correction for the number of tested diversity measures (*q* = 0.05/4 = 0.0125) or genera (*q =* 0.05/82 *=* 6.1×10^−4^). Initially,regressions were computed without controlling for covariates. Subsequently, significant (*q*<.05) bivariate associations of diversity measures or specific genera with the WPPSI-III FIQ score were explored by adding potential confounding variables as covariates in the regression model if they were associated with the respective diversity measure or genus at the nominal level (*p*<.05, same variables as in the REML models). Additional regression analyses were performed to test the association between the taxa and pathways identified by metagenomic sequencing and the WPPSI-III subscales. Additionally, we explored whether the maternal perceived stress during pregnancy related to the main findings, by testing the association of the PSS with the associated microbiome measures. Subjects with missing data for a WPPSI-III scale or covariates were excluded from the respective analyses (pairwise deletion).

#### Mediation analysis

Mediation analysis was used to investigate the hypothesis that the occurrence of an identified taxa mediates the effect of the most strongly associated covariates (i.e. smoking during pregnancy or current maternal smoking, see results) on the WPPSI-III full-scale IQ score, using quasi-Bayesian Monte Carlo simulation with 10,000 simulations as implemented in the mediation package.

## Results

### Study population

Microflora profiling was successful in 364 out of 380 samples (Figure 1). For the subsequent statistical analyses one subject with an outlying gestational age, 35 subjects with a lack of valid measures for any of the WPPSI-III scales, and 5 with irregularities in shipping/storage were excluded, resulting in a total of 323 subjects. For the remaining families, demographics (maternal age, child age, sex), pre-, peri- and postnatal factors (gestational age, birth weight, smoking, C-section, breastfeeding, maternal education, multilingual upbringing, child’s BMI, antibiotics use), and cognition (WPSSI-III FIQ, VIQ, PIQ, GLC) are depicted in Table 1.

**Table 1:** Information on pre-, peri-, and postnatal factors and cognition in the final study sample (n = 323). WPPSI-III = Wechsler Preschool and Primary Scale of Intelligence; SD = standard deviation; FIQ = full-scale IQ; GLC = general language composite; PIQ = performance IQ; VIQ = verbal IQ

|  | Frequency / Mean (SD) | N |
| --- | --- | --- |
| **Maternal age (years)** | 31.33 (4.89) | 323 |
| **Smoking during pregnancy** | 20.7% | 323 |
| **Gestational age (weeks)** | 39.14 (1.25) | 318 |
| **Birth weight (gram)** | 3,392 (468.48) | 317 |
| **Current maternal smoking** | 24.1% | 323 |
| **C-section** | 31.3% | 323 |
| **Breastfeeding** | 83.2% | 322 |
| **Sex (female)** | 53.2% | 323 |
| **Antibiotics last 6 months** | 31.4% | 322 |
| **Maternal education (years)** | 14.78 (2.23) | 323 |
| **Multilingual upbringing** | 23.4% | 321 |
| **Child age (months)** | 45.02 (1.00) | 323 |
| **Child BMI** | 15.44 (1.35) | 320 |
| **Perceived stress pregnancy** | 21.12 (8.36) | 240 |
| **Current perceived stress** | 23.91 (8.02) | 320 |
| **Shipment** | 18% | 323 |
| **WPPSI-III FIQ** | 103.30 (11.81) | 308 |
| **WPPSI-III VIQ** | 104.81 (11.72) | 311 |
| **WPPSI-III PIQ** | 100.62 (13.48) | 315 |
| **WPPSI-III GLC** | 102.33 (11.39) | 320 |

**Figure 1:**
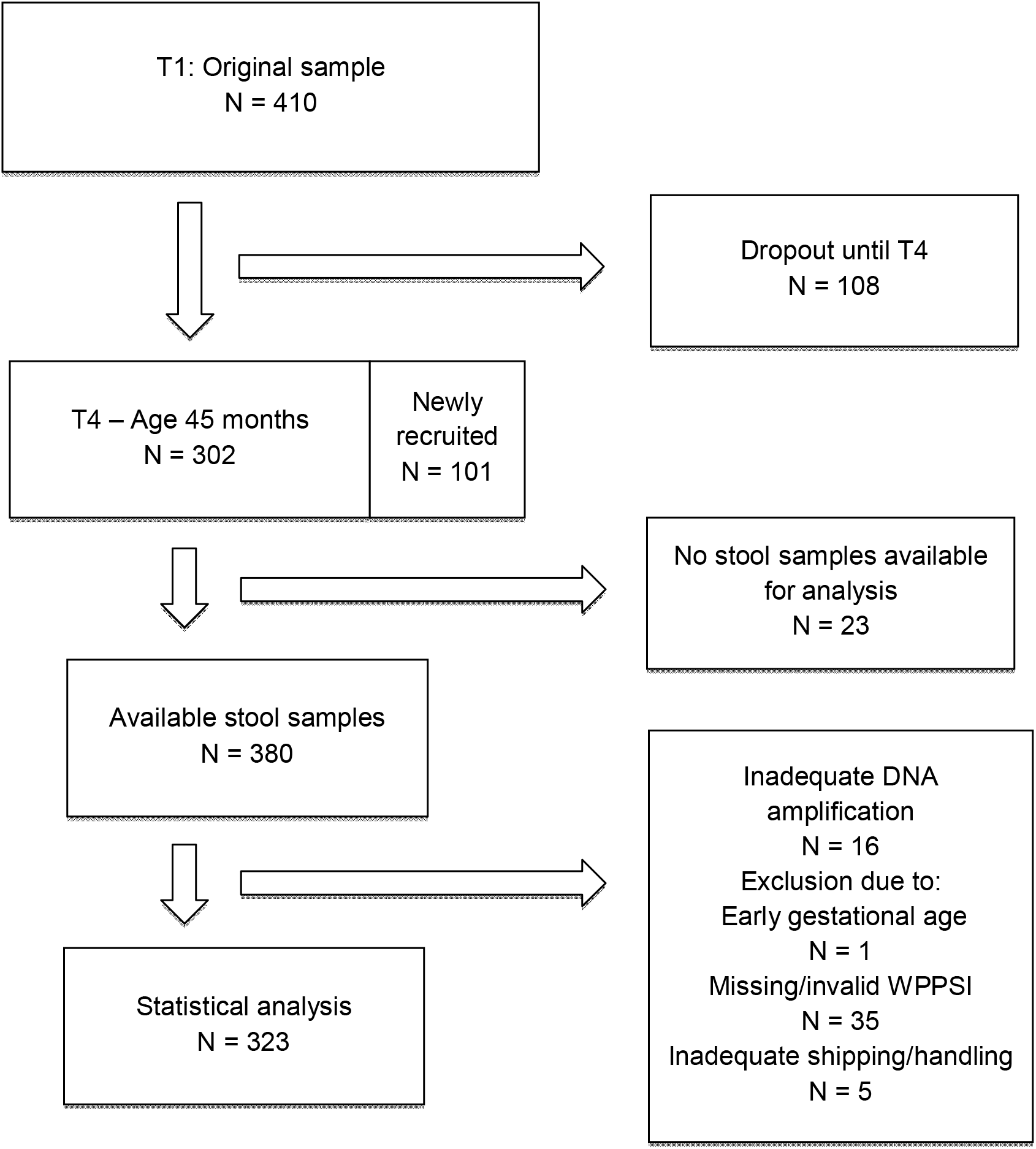
Consort flow diagram indicating the number of subjects participating in the POSEIDON study at the T1 and T4 time points, along with the number of subjects who provided stool samples and the number of samples that were included in the statistical analysis following successful library preparation and microbiome analyses.

### Variance in cognition explained by the microbiome

After quality and content filtering (removing environmental contaminants and keeping genera that present at least in 10 samples), 82 genera remained and were utilized in subsequent downstream analyzes. The variance explained in the WPPSI-III scales by the overall microbiome was characterized by restricted maximum likelihood (REML) models. These analyses showed that a significant proportion of the variance of WPPSI-III FIQ (variance explained = 13.3%, p=.0010) was association with the overall composition of the microbiome. Furthermore, the microbiome was associated with the subscales WPPSI-III VIQ and WPPSI-III GLC (variance explained = 15.8%; 10.3%, p=.000027; p=.0014 respectively) but not of WPPSI-III PIQ (0.5%; p = .46; Table 2). Of the assessed covariates, smoking during pregnancy, current smoking, breastfeeding, maternal education, multilingual upbringing, and shipment showed a significant association with the microbiome profile in REML models (see Supplementary Table S3). When those variables were jointly added as covariates to the REML cognition models, the explained variance of the WPPSI-III scales by variation in the overall composition of the microbiome variables was substantially reduced, with only the WPPSI-III VIQ scale remaining significant (see Supplementary Table S4). When each of these covariates was added individually to the REML model of WPPSI-III FIQ, the strongest reduction in explained variance was observed for smoking during pregnancy (see Supplementary Table S5).

**Table 2:** Explained variance of cognition phenotypes by variation in the microbiome estimated using restricted maximum likelihood (REML) models. (V/G)Vp = explained variance; CI = 95% confidence interval; WPPSI-III = Wechsler Preschool and Primary Scale of Intelligence Version III; FIQ = full-scale IQ; GLC = general language composite; PIQ = performance IQ; VIQ = verbal IQ.

| Phenotype | (V/G)Vp (CI) | *P* |
| --- | --- | --- |
| WPPSI-III FIQ | 13.4% (4.9%; 25.1%) | .0010 |
| WPPSI-III VIQ | 15.8% (7.1%; 29.4%) | .0000273 |
| WPPSI-III PIQ | 0.5% (0.0%; 12.4%) | .46 |
| WPPSI-III GLC | 10.3% (2.7%; 22.2%) | .0014 |

Significant negative correlations were observed between alpha diversity metrics and cognition. As shown in Table 3, the associations of WPPSI-III FIQ with Faith’s phylogenetic diversity index remained significant after correction for multiple testing. We observed significant (p<0.05) negative correlations of antibiotic use with Shannon diversity index and Pileou’s Evenness and of gestation age with Shannon diversity index and number of observed species, and a positive correlation of C-section with Faith’s phylogenetic diversity index (see Supplementary Figure S1). When we adjusted for these covariates in the association analyses with the respective diversity measures, we observed similar results, however the associations did not remain significant after correction for multiple testing (Supplementary Table S2).

**Table 3:**
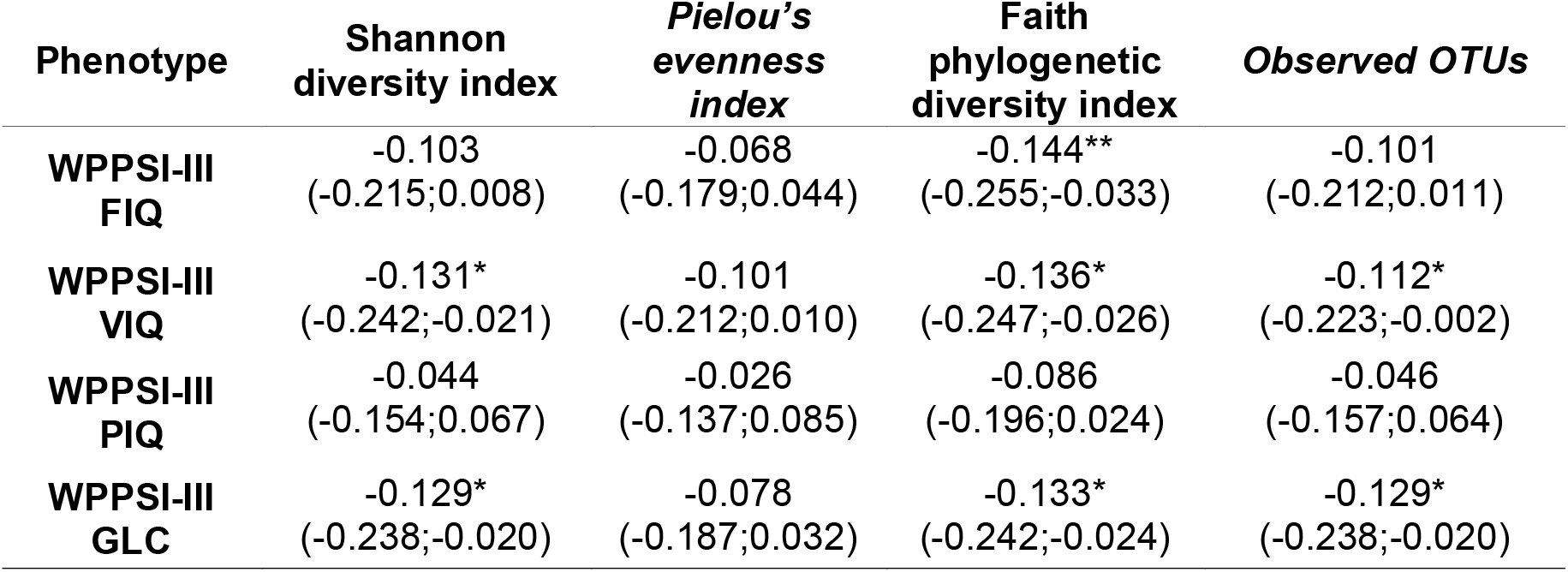
Associations of measures of cognition with measures of alpha diversity. Standardized betas and the 95% confidence interval are given; WPPSI-III = Wechsler Preschool and Primary Scale of Intelligence Version III; FIQ = full-scale IQ; GLC = general language composite; PIQ = performance IQ; VIQ = verbal IQ; * = *p*<.05, ** = p < 0.0125 (0.05 / 4)

As shown in Figures 2 and 3, relative abundance of one genus - an unidentified genus within the family *Enterobacteriaceae* were significantly associated with lower WPPSI-III FIQ (*ß*=-.20 [-.31; -0.095], *p*=.00031, *q*=.025, Supplementary Figure S2) after correction for the 82 tested genera. Relative abundance of this genus was also associated with lower performance on the WPPSI-III VIQ (*ß*=-19 [-.30; -.083] *p=*. 0.000636, *q*=.052), PIQ (*ß*=-.14 [-.25; -.031], *p*=.012, *q*=1), and GLC (*ß*=-.21 [-.32; -.10], *p*=.00015, *q*=.012) scores (Figure 2 A-D). Explorative analyses revealed an additional genus, Eubacterium genus of the Erysipelotrichaceae family (*ß*=-.20 [-.31; -.094], *p*=.0003047; *q*=.025) being associated with WPPSI-III verbal IQ subscales (*q*<.05).

**Figure 2:**
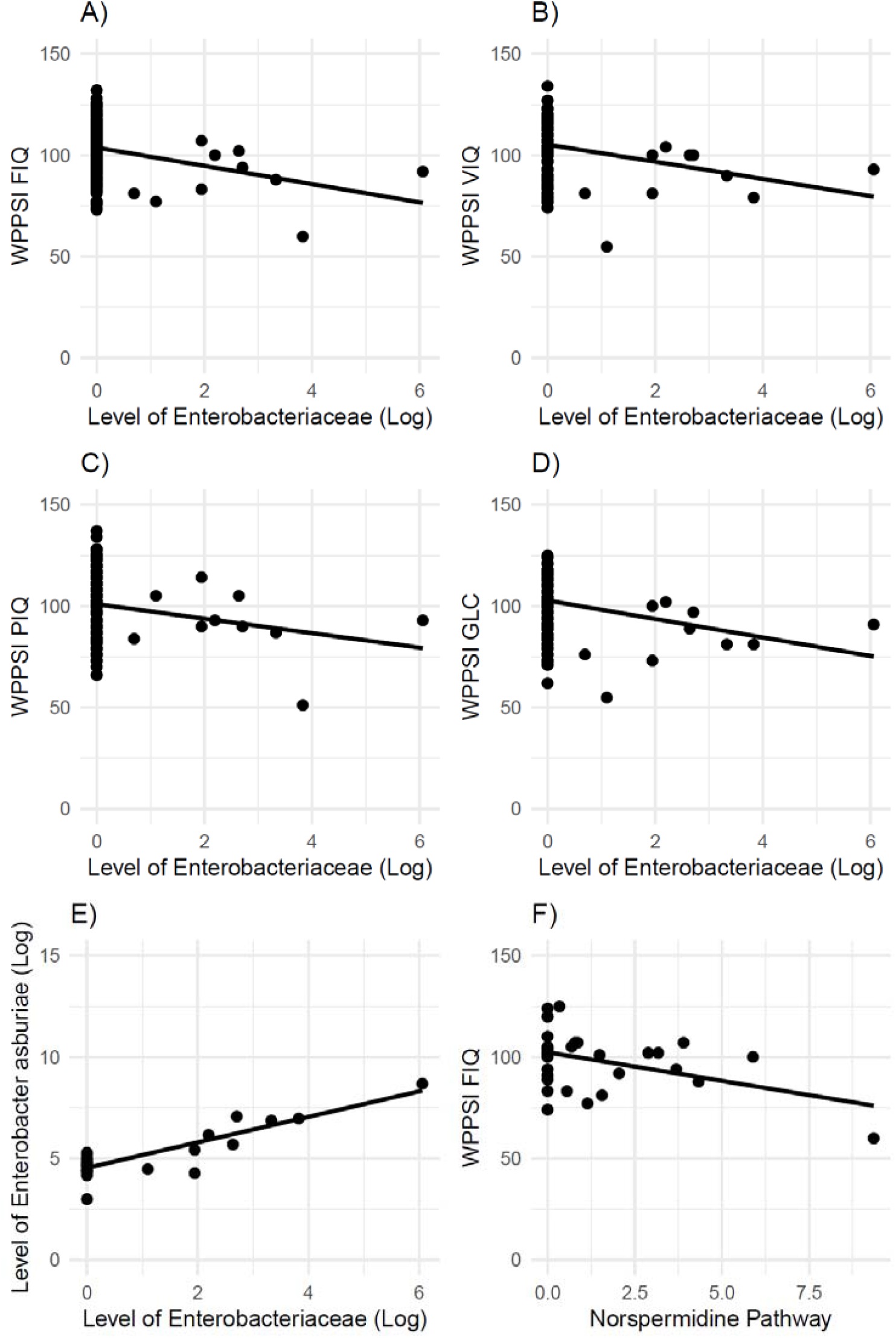
Correlation between the log transformed relative abundance of an unidentified genus within the family *Enterobacteriaceae* and A) the full scale IQ and B-D) subscales of the WPPSI-III, E) log transformed relative abundance of genus within the family *Enterobacteriaceae* with level of *Enterobacter asburiae* and F) WPPSI-III full-scale IQ score and predicted level of norspermidine biosynthesis. WPPSI-III = Wechsler Preschool and Primary Scale of Intelligence; FIQ = full-scale IQ; GLC = general language composite; PIQ = performance IQ; VIQ = verbal IQ

**Figure 3:**
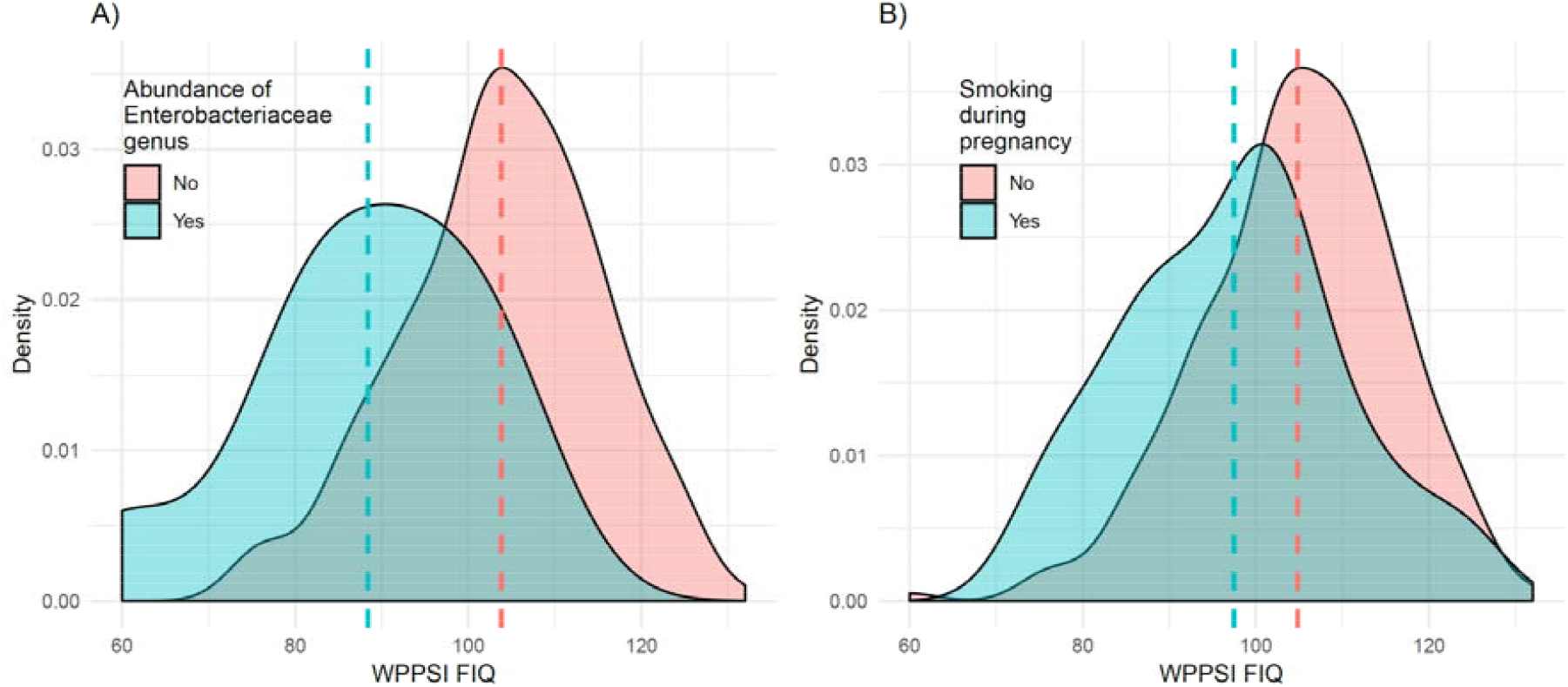
Distribution and mean of the full-scale WPPSI-III score A) by abundance of the genus within the family *Enterobacteriacea*e and B) by smoking during pregnancy. The group means are indicated by the dashed lines. WPPSI-III = Wechsler Preschool and Primary Scale of Intelligence Version III. FIQ = full-scale IQ

We subsequently carried out additional analyses with the unidentified genus within the family *Enterobacteriaceae*. It was nominally associated with following covariates (p<.05): maternal age (*ß*=.13 [-.24; -.024], *p*=.018), shipment (*ß*=-.18 [.070; .28], *p*=.0014), and more pronounced with maternal cigarette smoking during pregnancy (*ß*=.22 [.12; .33], *p*=0.0000505, Figure 3B) and smoking at T4 (*ß*=.18 [.077; .29], *p*=0.00088). The relationship between the unidentified genus within the family *Enterobacteriaceae* and WPPSI-III FIQ remained nominally significant when those variables were employed as covariates (*ß*=-.14 [-.25; -.026], *p*=.016, *q* = 1) in a regression model. As the variables most strongly associated with the genus were maternal smoking during pregnancy and current (T4) maternal smoking, those were further explored in mediation models. Mediation analyses (Fig 3.) revealed a significant mediation effect of this genus in the family Enterobacteriaceae for the effects of maternal smoking during pregnancy (proportion mediated = 13.7% [3.4%; 32%]; *p*=.007) and current smoking (proportion mediated = 12.0% [3.1%; 29%]; p=.0034) on WPPSI-III FIQ (controlling for age of the mother and shipment). Maternal smoking during T4 and maternal smoking during pregnancy were strongly associated (r = .64, p < 0.001), and a joint model predicting *Enterobacteriaceae* levels with both smoking variables indicated significant effects of pregnancy smoking (*ß*=-.18 [.039; .33], *p*=.013) but not for current smoking (*ß*=-.071 [-.072; .21], *p*=.33).

We further characterized the microbiome by metagenomic analysis performed in a subset of 33 samples (demographics details of subset are shown in Supplemental Table S1C). This analysis indicated that the organisms which belongs to this unidentified genus of the family *Enterobacteriaceae* were most closely related to *Enterobacter asburiae* (*ß*=.87 [.70; 1.0], *p*=3.8×10^−11^, Figure 2E), and were also related to *Enterobacter cloacae* (*ß*=.76 [.54; .99], *p*=2.4×10^−7^) and *Kluyvera intermedia* (*ß*=.51 [.20; .81], *p*=.0026). Of those organism, *Enterobacter cloacae* was significantly associated with the WPPSI-III FIQ, and *Enterobacter cloacae and Enterobacter asburiae* were associated with the WPPSI-III PIQ subscale. In the subjects for which the data was available (T1: n=240, T4: n=320), we additionally explored whether the associations of WPPSI FIQ with Faith’s phylogenetic diversity index and the Enterobacteriaceae genus were affected by perceived stress during pregnancy and current perceived stress. The PSS at T1 was not significantly associated with Faith’s phylogenetic diversity index (r=-.11; *p*=.074) but showed a nominal association with the Enterobacteriaceae genus species (r=.15; *p*=.022). The PSS at T4 was neither associated with the genus nor Faith’s phylogenetic diversity metric (p>.31). When PSS T1 was added as an additional predictor to the model with maternal age, shipment, smoking during pregnancy, and smoking at T4, the relationship between the unidentified genus within the family Enterobacteriaceae and WPPSI-III FIQ remained nominally significant (ß=-.16 [-.29; -.032], *p*=.015).

We also employed metabolic profiling to explore the relationship between cognitive functioning and the metabolic potential of the microbiome explored by metagenomic sequencing (details of metagenomics sequencing subset shown in Supplementary Table S1C). We found a nominally significant negative association between the WPPSI-III FIQ and the predicted level of norspermidine after adjusting for the covariates smoking during pregnancy, present cigarette smoking, maternal age, and shipment (*ß*=-.47 [-.82; -.12], *p*=.014; Figure 2F; Details of the components of this pathway are shown in Supplementary Figure S3, all other pathways p > 0.05).

## Discussion

The present prospective cohort study revealed an association between the composition of the fecal bacterial microflora and cognitive functioning as measured by the WPPSI-III in healthy children at 45 months of age.

Overall, we found that a significant portion of the difference in cognitive functioning of these children as measured by the WPPSI-III is correlated with variance in the composition of their fecal microflora. Of interest, this correlation was observed for the FIQ and the language-related subscales (VIQ & GLC), but not for the PIQ subscale of the WPPSI-III. Additionally, we found evidence of a negative association of cognition measures with alpha diversity, with a higher diversity being associated with lower cognitive scores. Again, the significant associations were limited to the FIQ and the language-related subscales (VIQ & GLC). Such associations between the composition of the bacterial microflora and cognitive functioning are consistent with the findings of Carlson and colleagues (28). They measured the fecal microbiome in children at the age of one and assessed cognitive functioning (Mullen scales of early learning) at the age of one and two years. They identified three distinct bacterial clusters, where significant differences between the clusters were associated with cognitive functioning as measured by Mullen scales of early learning by both overall score and by subdomain performance at the age of two years. As observed in the current study, they found negative correlations in alpha diversity measures, with higher diversity being associated with lower cognitive functioning scores, however, no association remained significant after correction for multiple testing.

At the genus level, we found that lower levels of cognitive functioning in preschool-aged children were associated with an increased level of DNA of an unidentified genus of the family Enterobacteriaceae. In previous studies, supportive evidence links exposure to similar organisms to altered cognitive functioning in aging humans (62) and in animal models of aging (63). Notably, antibiotics usage, which has been linked to lower cognitive abilities later in childhood (33) showed no correlation with this genus. The mechanisms of these associations are still unidentified but are likely to involve alterations of immune activity within the brain via alterations of the gut-immune-brain axis (64,65). Metagenomic sequencing of a subset of samples indicated that the associated taxon was closely related to *Enterobacter asburiae* and *Enterobacter cloacae*. These are highly homologous organisms which inhabit the gastrointestinal tract of infants and young children and are occasionally associated with symptomatic infections (66,67). It is yet to be clarified whether this taxon represents a novel species related to both *Enterobacter asburiae* and *Enterobacter cloacae* or a mixture of several related species within the *Enterobacter cloacae* complex (68). Additional metagenomic sequencing will be required to further characterize these organisms and determined mechanisms by which they might be related to cognitive functioning.

The finding of a potential association between lower levels of cognitive functioning and norspermidine synthesis is also of interest. Norspermidine is a polyamine generated by bacteria that is structurally and functionally similar to spermidine, a polyamine synthesized in humans and other animals (69). Spermidine and other polycationic polyamines have been shown to improve cognitive functioning in animals and are being investigated for the prevention of cognitive decline in aging humans (70). The mechanism of action is uncertain but likely to be related to the biological functions of polycationic molecules, including modulation of the activity of ionic channels, protein synthesis, protein kinases, and cell proliferation and apoptosis. (71). Loss of function mutations in spermidine synthase are associated with decreased levels of cognitive functioning in Snyder-Robinson syndrome (72). Animal model studies indicate that the mechanism of the cognitive dysfunction in this syndrome is likely related to lysosomal dysfunction and oxidative stress (73). However, as these analyses were based on a comparatively small subset of the cohort, and this association only reached nominal significance (p < 0.05), additional studies will be required to conform this potential association and to define precise mechanisms of association.

Previous studies have indicated that cognition in early childhood is impaired by maternal smoking (74). The results of the mediation analyses in the present study indicate that the effects of both smoking during pregnancy and current smoking on cognition might be (partially) mediated by the microbiome. Maternal smoking has been linked to adverse outcomes in both somatic and mental health of the offspring (75). Both animal and human studies indicate possible effects of smoking on the microbiome. For example, in rats, smoking was associated with reductions in *Bifidobacteria* (76), and side-stream smoking increased the levels of *Clostridium* and reduced the levels of *Firmicutes* (*Lactococci* and *Reminococcus*) and *Enterobacteriacae* families in mice (77). In humans, it has been shown, that smokers and non-smokers differ in regard to their microbiome (e.g. 78,79) and that changes in smoking behavior affect the microbiome (80). As the mother’s microbiome has been shown to influence the microbiome of the child (15), it is plausible that cigarette smoking during pregnancy could influence the child’s microbiome profile as well. Notably, prenatal smoking showed a stronger link to the *Enterobacteriaceae* genus than current smoking in the present study.

There are several possible caveats to our findings. The significant correlation between altered level of this unidentified genus of the Enterobacteriaceae family and cognitive functioning might be due to a direct connection between the microbiome and brain development. However, it is also possible that, the presence of this genus might be related to unmeasured concomitant factors, such as altered diet, family structure, and social interactions (62). Among the above listed concomitant factors, diet is quite unlikely, since all children except one were attending national daycare where they have been exposed to a very similar diet. Several postnatal factors were not investigated in the present study, including household size, exposure to pets and other animals, travel, urban vs. suburban living, and additional lifestyle factors (e.g. diet or parental alcohol consumption), which might be associated with the microbiome, smoking and cognitive functioning. The present study indicates that such covariates can influence observed associations of outcomes with the microbiome. The potential role of these factors as mediators between the microbiome and cognitive development should be the subject of additional studies. While we did not observe a strong influence of measures of prenatal and current perceived stress on the association of the microbiome with cognition, the direct associations of pre-, peri- and postnatal stress measures with the microbiome should be assessed in more detail in the future. Another limitation of this present study was that in the case of the subset of subjects who were newly recruited at T4, environmental factors from earlier time points such as smoking during pregnancy or breastfeeding could only be assessed retrospectively by self-report. However, these variables were taken into account as binary variable in our analyses, hence the retrospective self-report is unlikely to introduce strong bias. Another limitation of our study is that our microbial data collection is cross-sectional, precluding studies of temporal variation. Additional longitudinal studies are planned to confirm our findings

Our study documents an association between intestinal microflora composition and cognitive functioning, with the microbiome mediating the effect of maternal smoking on cognition in a group of otherwise healthy preschool-aged children. The microbiome of a child can be altered by environmental factors, such as diet, antibiotics, and exposure to cigarette smoke, as well as therapeutically by administration of probiotic or prebiotic medications (81). Follow-up studies need to test the possibility of a direct interaction between the microbiome and cognitive functioning in this age group. The establishment of a cause-and-effect association may lead to the evaluation of interventions directed at improving cognitive functioning in infants and young children.

## Data Availability

The datasets generated during and/or analyzed during the current study are not publicly available due to privacy regulations but are available from the authors on reasonable request.

## Acknowledgements

We thank all parents and children for taking part in this study and our student employees and interns for their support with data acquisition and data entry. We thank Regev Schweiger (FIESTA) and Jian Yang (OSCA) for feedback on the application of their software.

