## Supplementary Material for "Microbiome profiles are associated with cognitive functioning in 45-month-old children"

[1]Department of Genetic Epidemiology in Psychiatry, Central Institute of Mental Health, Medical Faculty Mannheim, University of Heidelberg, Mannheim, Germany; [2]Department of Psychiatry and Psychotherapy, Central Institute of Mental Health, Medical Faculty Mannheim, University of Heidelberg, Mannheim, Germany; [3] Stanley Division of Developmental Neurovirology, Department of Pediatrics. Johns Hopkins School of Medicine, Baltimore, MD, United States of America

*contributed equally

| | Frequency / Mean (SD) | N |
|---|---|---|
| **Maternal age (years)** | 31.76 (4.77) | 244 |
| **Smoking during pregnancy** | 18.4% | 244 |
| **Gestational age (weeks)** | 39.22 (1.18) | 244 |
| **Current maternal smoking** | 22.5% | 244 |
| **Birth weight (grams)** | 3428.64 (459.01) | 244 |
| **C-section** | 26.2% | 244 |
| **Breastfeeding** | 86.4% | 243 |
| **Sex (female)** | 56.1% | 244 |
| **Antibiotics last 6 months** | 29.2% | 243 |
| **Maternal education (years)** | 15.09 (2.17) | 244 |
| **Multilingual** | 22.7% | 242 |
| **Child age (months)** | 44.85 (0.91) | 244 |
| **Child BMI** | 15.39 (1.32) | 241 |
| **Perceived stress pregnancy** | 21.12 (8.36) | 240 |
| **Current perceived stress** | 23.72 (8.15) | 241 |
| **Shipment** | 16.4% | 244 |
| **WPPSI-III FIQ** | 104.27 (10.96) | 231 |
| **WPPSI-III VIQ** | 105.75 (10.57) | 234 |
| **WPPSI-III PIQ** | 101.44 (13.11) | 236 |
| **WPPSI-III GLC** | 103.56 (10.14) | 241 |

**Supplementary Table S1A: Information on pre-, peri-, and postnatal factors and cognition in the original study sample (n = 244).** WPPSI-III = Wechsler Preschool and Primary Scale of Intelligence; SD = standard deviation; FIQ = full scale IQ; GLC = general language composite; PIQ = performance IQ; VIQ = verbal IQ

| | Frequency / Mean (SD) | N |
|---|---|---|
| **Maternal age (years)** | 29.99 (5.05) | 79 |
| **Smoking during pregnancy** | 27.9% | 79 |
| **Gestational age (weeks)** | 38.91 (1.44) | 74 |
| **Birth weight (grams)** | 3269.52 (482.07) | 73 |
| **Current maternal smoking** | 29.1% | 79 |
| **C-section** | 46.8% | 79 |
| **Breastfeeding** | 73.4% | 79 |
| **Sex (female)** | 55.7% | 79 |
| **Antibiotics last 6 months** | 38.0% | 79 |
| **Maternal education (years)** | 13.84 (2.14) | 79 |
| **Multilingual** | 25.3% | 79 |
| **Child age (months)** | 45.54 (1.07) | 79 |
| **Child BMI** | 15.59 (1.42) | 79 |
| **Current perceived stress** | 24.49 (7.64) | 79 |
| **Shipment** | 22.8% | 79 |
| **WPPSI-III FIQ** | 100.39 (13.97) | 77 |
| **WPPSI-III VIQ** | 101.96 (14.39) | 77 |
| **WPPSI-III PIQ** | 98.15 (14.35) | 79 |
| **WPPSI-III GLC** | 98.59 (13.99) | 79 |

**Supplementary Table S1B: Information on pre-, peri-, and postnatal factors and cognition in the subsample newly recruited at T4 (n = 79).** WPPSI-III = Wechsler Preschool and Primary Scale of Intelligence; SD = standard deviation; FIQ = full scale IQ; GLC = general language composite; PIQ = performance IQ; VIQ = verbal IQ

| | Frequency / Mean (SD) | N |
|---|---|---|
| **Maternal age (years)** | 29.12 (6.01) | 33 |
| **Smoking during pregnancy** | 48.5% | 33 |
| **Gestational age (weeks)** | 39.00 (1.61) | 31 |
| **Birth weight (grams)** | 3213.74 (449.45) | 31 |
| **Current maternal smoking** | 51.5% | 33 |
| **C-section** | 57.6% | 33 |
| **Breastfeeding** | 51.5% | 33 |
| **Sex (female)** | 69.7% | 33 |
| **Antibiotics last 6 months** | 48.5% | 33 |
| **Maternal education (years)** | 13.55 (2.45) | 33 |
| **Child age (months)** | 45.09 (1.04) | 33 |
| **Child BMI** | 15.32 (1.36) | 33 |
| **Shipment** | 27.3% | 33 |
| **WPPSI-III FIQ** | 98.67 (14.27) | 33 |
| **WPPSI-III VIQ** | 98.61 (13.40) | 33 |
| **WPPSI-III PIQ** | 98.91 (16.27) | 33 |
| **WPPSI-III GLC** | 96.21 (13.98) | 33 |

**Supplementary Table S1C: Information on pre-, peri-, and postnatal factors and cognition in the subset sample in which metagenomic sequencing data was analysed**. WPPSI-III = Wechsler Preschool and Primary Scale of Intelligence; FIQ = full scale IQ; GLC = general language composite; PIQ = performance IQ; VIQ = verbal IQ

| Phenotype | Shannon diversity index | *Pielou's evenness index* | Faith phylogenetic diversity index | *Observed OTUs* |
|---|---|---|---|---|
| **WPPSI-III FIQ** | -0.093 (-0.204;0.018) | -0.071 (-0.182;0.04) | -0.132* (-0.243;-0.021) | -0.086 (-0.197;0.025) |
| **WPPSI-III VIQ** | -0.106 (-0.217;0.004) | -0.097 (-0.207;0.014) | -0.123* (-0.234;-0.012) | -0.088 (-0.199;0.023) |
| **WPPSI-III PIQ** | -0.054 (-0.164;0.057) | -0.035 (-0.145;0.075) | -0.078 (-0.188;0.032) | -0.05 (-0.16;0.06) |
| **WPPSI-III GLC** | -0.104 (-0.214;0.005) | -0.076 (-0.185;0.032) | -0.118* (-0.228;-0.009) | -0.10 (-0.21;0.009) |

**Supplementary Table S2: Associations of measures of cognition with measures of alpha diversity while controlling for covariates associated with alpha diversity measures.** Standardized betas and the 95% confidence interval are given; WPPSI-III = Wechsler Preschool and Primary Scale of Intelligence; FIQ = full scale IQ; GLC = general language composite; PIQ = performance IQ; VIQ = verbal IQ; * = *p*<.05

| Variable | (V/G)Vp | *P* |
|---|---|---|
| Maternal age | 0.39% | 0.4528 |
| Smoking during pregnancy | 11.75% | 0.0014 |
| Gestational age | 0.00% | 0.5000 |
| Birth weight | 0.89% | 0.3782 |
| Current maternal smoking | 8.82% | 0.0035 |
| C-section | 4.41% | 0.1297 |
| Breastfeeding | 9.58% | 0.0021 |
| Sex (female) | 3.90% | 0.1168 |
| Antibiotics last 6 months | 4.11% | 0.2177 |
| Maternal education | 11.94% | 0.0004 |
| Multilingual upbringing | 9.38% | 0.0178 |
| Child age | 2.34% | 0.2579 |
| Child BMI | 0.00% | 0.5000 |
| Shipment | 18.97% | 0.0000 |

**Supplementary Table S3: Explained variance in potential confounding variables by variation in the microbiome estimated using restricted maximum likelihood (REML) models.** (V/G)Vp = explained variance

| Phenotype | No covariates (V/G)Vp (CI) | *P* | With covariates[#] (V/G)Vp | *P* |
|---|---|---|---|---|
| **WPPSI-III FIQ** | 13.4% (4.9%; 25.1%) | .0010 | 1.6% | .40 |
| **WPPSI-III VIQ** | 15.8% (7.1%; 29.4%) | .0000273 | 9.4% | .024 |
| **WPPSI-III PIQ** | 0.5% (0.0%; 12.4%) | .46 | <0.01% | .50 |
| **WPPSI-III GLC** | 10.3% (2.7%; 22.2%) | .0014 | 2.8% | .21 |

**Supplementary Table S4: Explained variance of cognition phenotypes by variation in the microbiome estimated using restricted maximum likelihood (REML) models.** (V/G)Vp = explained variance; CI = 95% confidence interval; WPPSI-III = Wechsler Preschool and Primary Scale of Intelligence Version III; FIQ = full-scale IQ; GLC = general language composite; PIQ = performance IQ; VIQ = verbal IQ. [#]Covariates were added to the models when they showed a significant association with the microbiome in univariate REML models: maternal education, breastfeeding, smoking during pregnancy, current smoking, multilingual upbringing, shipment

| **WPPSI-III FIQ** | (V/G)Vp | *P* |
|---|---|---|
| **No covariates** | 13.4% | .0010 |
| **Pregnancy smoking** | 5.9% | .088 |
| **Current smoking** | 8.1% | .043 |
| **Breastfeeding** | 11.3% | .0093 |
| **Maternal education** | 8.4% | .049 |
| **Multilingual upbringing** | 13.0% | .0018 |
| **Shipment** | 13.1% | .0014 |
| **All covariates** | 1.6% | .40 |

**Supplementary Table S5: Explained variance of WPPSI-III FIQ by variation in the microbiome estimated using restricted maximum likelihood (REML) models, depending on the covariates in the REML models.** (V/G)Vp = explained variance; WPPSI-III = Wechsler Preschool and Primary Scale of Intelligence Version III; FIQ = full-scale IQ.

| Pathway | Description |
|---|---|
| 3-HYDROXYPHENYLACETATE-DEGRADATION-PWY | 4-hydroxyphenylacetate degradation |
| AEROBACTINSYN-PWY | aerobactin biosynthesis |
| ALLANTOINDEG-PWY | superpathway of allantoin degradation in yeast |
| ARGDEG-PWY | superpathway of L-arginine, putrescine, and 4-aminobutanoate degradation |
| AST-PWY | L-arginine degradation II (AST pathway) |
| CATECHOL-ORTHO-CLEAVAGE-PWY | catechol degradation to β-ketoadipate |
| ECASYN-PWY | enterobacterial common antigen biosynthesis |
| ENTBACSYN-PWY | enterobactin biosynthesis |
| GLUCOSE1PMETAB-PWY | glucose and glucose-1-phosphate degradation |
| GLUDEG-II-PWY | L-glutamate degradation VII (to butanoate) |
| GLYCOCAT-PWY | glycogen degradation I (bacterial) |
| GLYCOL-GLYOXDEG-PWY | superpathway of glycol metabolism and degradation |
| GLYCOLYSIS-TCA-GLYOX-BYPASS | superpathway of glycolysis, pyruvate dehydrogenase, TCA, and glyoxylate bypass |
| GLYOXYLATE-BYPASS | glyoxylate cycle |
| HCAMHPDEG-PWY | 3-phenylpropanoate and 3-(3-hydroxyphenyl)propanoate degradation to 2-oxopent-4-enoate |
| KDO-NAGLIPASYN-PWY | superpathway of (Kdo)2-lipid A biosynthesis |
| KETOGLUCONMET-PWY | ketogluconate metabolism |
| LPSSYN-PWY | superpathway of lipopolysaccharide biosynthesis |
| METHGLYUT-PWY | superpathway of methylglyoxal degradation |
| NAGLIPASYN-PWY | lipid IVA biosynthesis |
| ORNARGDEG-PWY | superpathway of L-arginine and L-ornithine degradation |
| ORNDEG-PWY | superpathway of ornithine degradation |
| P105-PWY | TCA cycle IV (2-oxoglutarate decarboxylase) |
| P125-PWY | superpathway of (R,R)-butanediol biosynthesis |
| P162-PWY | L-glutamate degradation V (via hydroxyglutarate) |
| P163-PWY | L-lysine fermentation to acetate and butanoate |
| P23-PWY | reductive TCA cycle I |
| P241-PWY | coenzyme B biosynthesis |
| POLYAMINSYN3-PWY | superpathway of polyamine biosynthesis II |
| PROTOCATECHUATE-ORTHO-CLEAVAGE-PWY | protocatechuate degradation II (ortho-cleavage pathway) |
| PWY0-1241 | ADP-L-glycero-β-D-manno-heptose biosynthesis |
| PWY0-1261 | anhydromuropeptides recycling |
| PWY0-1277 | 3-phenylpropanoate and 3-(3-hydroxyphenyl)propanoate degradation |
| PWY0-1338 | polymyxin resistance |
| PWY0-1415 | superpathway of heme biosynthesis from uroporphyrinogen-III |
| PWY0-1479 | tRNA processing |
| PWY0-1533 | methylphosphonate degradation I |
| PWY0-321 | phenylacetate degradation I (aerobic) |

| | |
|---|---|
| PWY0-41 | allantoin degradation IV (anaerobic) |
| PWY0-42 | 2-methylcitrate cycle I |
| PWY-1861 | formaldehyde assimilation II (RuMP Cycle) |
| PWY-2201 | folate transformations I |
| PWY-2723 | trehalose degradation V |
| PWY-3781 | aerobic respiration I (cytochrome c) |
| PWY-4321 | L-glutamate degradation IV |
| PWY-4361 | S-methyl-5-thio-α-D-ribose 1-phosphate degradation |
| PWY-4702 | phytate degradation I |
| PWY-5005 | biotin biosynthesis II |
| PWY-5083 | NAD/NADH phosphorylation and dephosphorylation |
| PWY-5088 | L-glutamate degradation VIII (to propanoate) |
| PWY-5138 | unsaturated, even numbered fatty acid β-oxidation |
| PWY-5392 | reductive TCA cycle II |
| PWY-5417 | catechol degradation III (ortho-cleavage pathway) |
| PWY-5431 | aromatic compounds degradation via β-ketoadipate |
| PWY-561 | superpathway of glyoxylate cycle and fatty acid degradation |
| PWY-5656 | mannosylglycerate biosynthesis I |
| PWY-5675 | nitrate reduction V (assimilatory) |
| PWY-5692 | allantoin degradation to glyoxylate II |
| PWY-5705 | allantoin degradation to glyoxylate III |
| PWY-5723 | Rubisco shunt |
| PWY-5747 | 2-methylcitrate cycle II |
| PWY-5791 | 1,4-dihydroxy-2-naphthoate biosynthesis II (plants) |
| PWY-5837 | 1,4-dihydroxy-2-naphthoate biosynthesis I |
| PWY-5838 | superpathway of menaquinol-8 biosynthesis I |
| PWY-5840 | superpathway of menaquinol-7 biosynthesis |
| PWY-5855 | ubiquinol-7 biosynthesis (prokaryotic) |
| PWY-5856 | ubiquinol-9 biosynthesis (prokaryotic) |
| PWY-5857 | ubiquinol-10 biosynthesis (prokaryotic) |
| PWY-5861 | superpathway of demethylmenaquinol-8 biosynthesis |
| PWY-5863 | superpathway of phylloquinol biosynthesis |
| PWY-5897 | superpathway of menaquinol-11 biosynthesis |
| PWY-5898 | superpathway of menaquinol-12 biosynthesis |
| PWY-5899 | superpathway of menaquinol-13 biosynthesis |
| PWY-5910 | superpathway of geranylgeranyldiphosphate biosynthesis I (via mevalonate) |
| PWY-5920 | superpathway of heme biosynthesis from glycine |
| PWY-6060 | malonate degradation II (biotin-dependent) |
| PWY-6071 | superpathway of phenylethylamine degradation |
| PWY-6145 | superpathway of sialic acids and CMP-sialic acids biosynthesis |
| PWY-6165 | chorismate biosynthesis II (archaea) |
| PWY-622 | starch biosynthesis |
| PWY-6269 | adenosylcobalamin salvage from cobinamide II |

| | |
|---|---|
| PWY-6309 | L-tryptophan degradation XI (mammalian, via kynurenine) |
| PWY-6318 | L-phenylalanine degradation IV (mammalian, via side chain) |
| PWY-6344 | L-ornithine degradation II (Stickland reaction) |
| PWY-6383 | mono-trans, poly-cis decaprenyl phosphate biosynthesis |
| PWY-6467 | Kdo transfer to lipid IVA III (Chlamydia) |
| PWY-6470 | peptidoglycan biosynthesis V (β-lactam resistance) |
| PWY-6562 | norspermidine biosynthesis |
| PWY-6690 | cinnamate and 3-hydroxycinnamate degradation to 2-oxopent-4-enoate |
| PWY-6708 | ubiquinol-8 biosynthesis (prokaryotic) |
| PWY-6731 | starch degradation III |
| PWY-6803 | phosphatidylcholine acyl editing |
| PWY-6823 | molybdenum cofactor biosynthesis |
| PWY-6837 | fatty acid beta-oxidation V (unsaturated, odd number, di-isomerase dependent) |
| PWY-6891 | thiazole biosynthesis II (Bacillus) |
| PWY-6892 | thiazole biosynthesis I (E. coli) |
| PWY-6895 | superpathway of thiamin diphosphate biosynthesis II |
| PWY-6953 | dTDP-3-acetamido-3,6-dideoxy-α-D-galactose biosynthesis |
| PWY-7118 | chitin degradation to ethanol |
| PWY-7200 | superpathway of pyrimidine deoxyribonucleoside salvage |
| PWY-7204 | pyridoxal 5'-phosphate salvage II (plants) |
| PWY-7269 | NAD/NADP-NADH/NADPH mitochondrial interconversion (yeast) |
| PWY-7279 | aerobic respiration II (cytochrome c) (yeast) |
| PWY-7294 | xylose degradation IV |
| PWY-7312 | dTDP-D-β-fucofuranose biosynthesis |
| PWY-7316 | dTDP-N-acetylviosamine biosynthesis |
| PWY-7332 | superpathway of UDP-N-acetylglucosamine-derived O-antigen building blocks biosynthesis |
| PWY-7385 | 1,3-propanediol biosynthesis (engineered) |
| PWY-7409 | phospholipid remodeling (phosphatidylethanolamine, yeast) |
| PWY-7446 | sulfoglycolysis |
| PWY-7616 | methanol oxidation to carbon dioxide |
| PWY-821 | superpathway of sulfur amino acid biosynthesis (Saccharomyces cerevisiae) |
| PWY-822 | fructan biosynthesis |
| REDCITCYC | TCA cycle VIII (helicobacter) |
| RUMP-PWY | formaldehyde oxidation I |
| TCA-GLYOX-BYPASS | superpathway of glyoxylate bypass and TCA |
| TEICHOICACID-PWY | teichoic acid (poly-glycerol) biosynthesis |
| THISYN-PWY | superpathway of thiamin diphosphate biosynthesis I |
| THREOCAT-PWY | superpathway of L-threonine metabolism |
| UBISYN-PWY | superpathway of ubiquinol-8 biosynthesis (prokaryotic) |
| URDEGR-PWY | superpathway of allantoin degradation in plants |
| VALDEG-PWY | L-valine degradation I |

**Supplementary Table S1C: List of tested metabolic pathways**

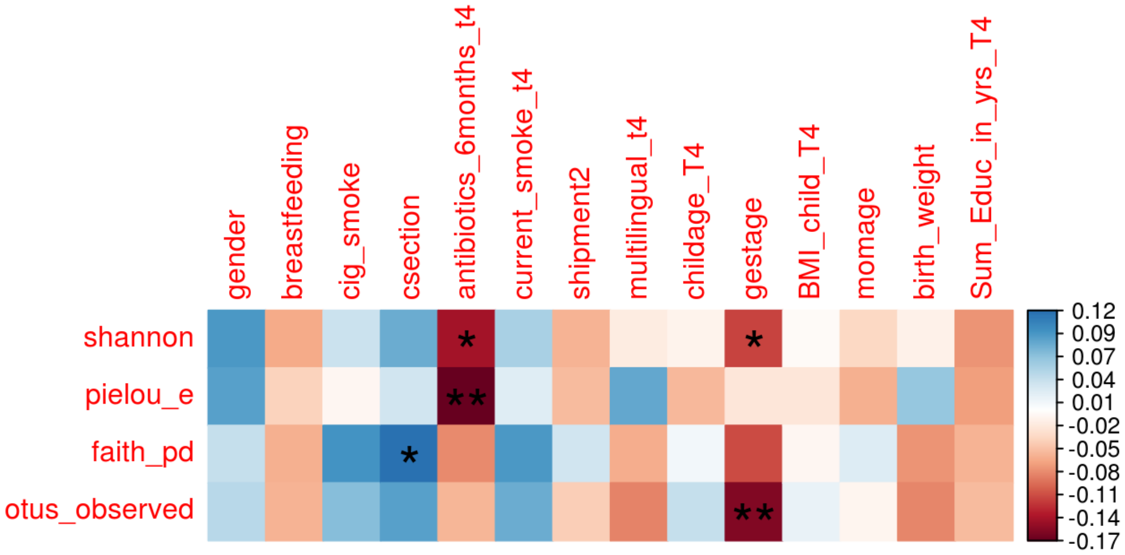

**Supplementary Figure S1: Correlation plot of potential confounders with alpha diversity measures.** * $p < 0.05$; * $p < 0.01$

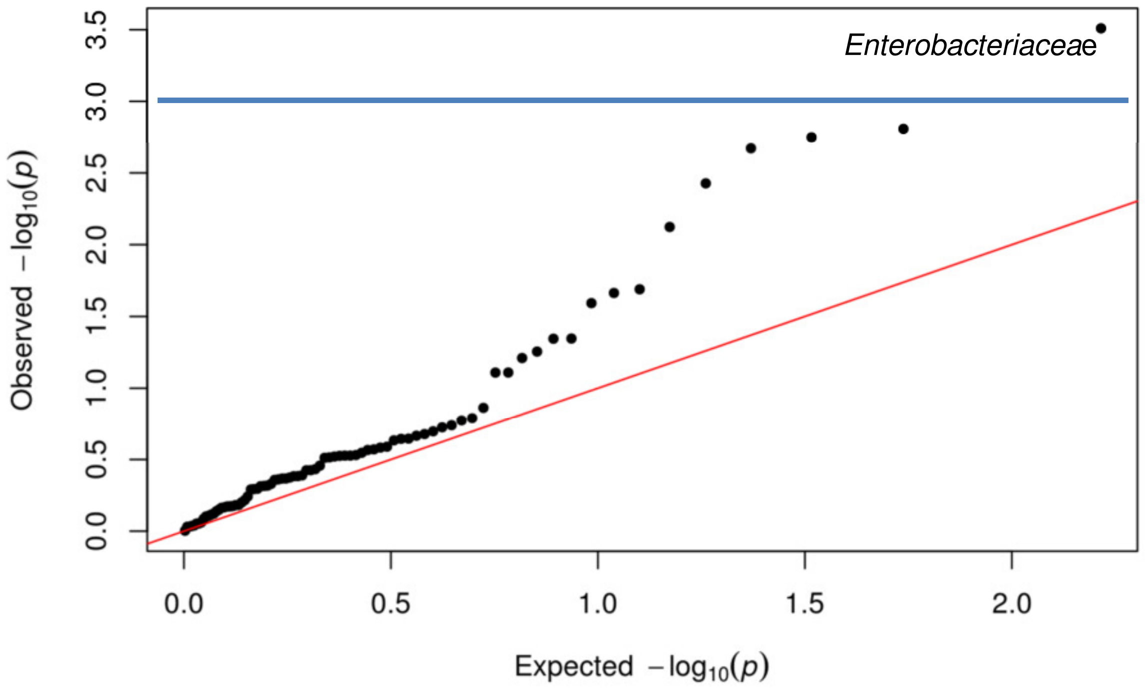

**Supplementary Figure S2: QQ plot of Single taxon associations with WPPSI-III full-scale IQ score.** The blue line indicates the level of significance after Bonferroni correction (0.05/82 genera).

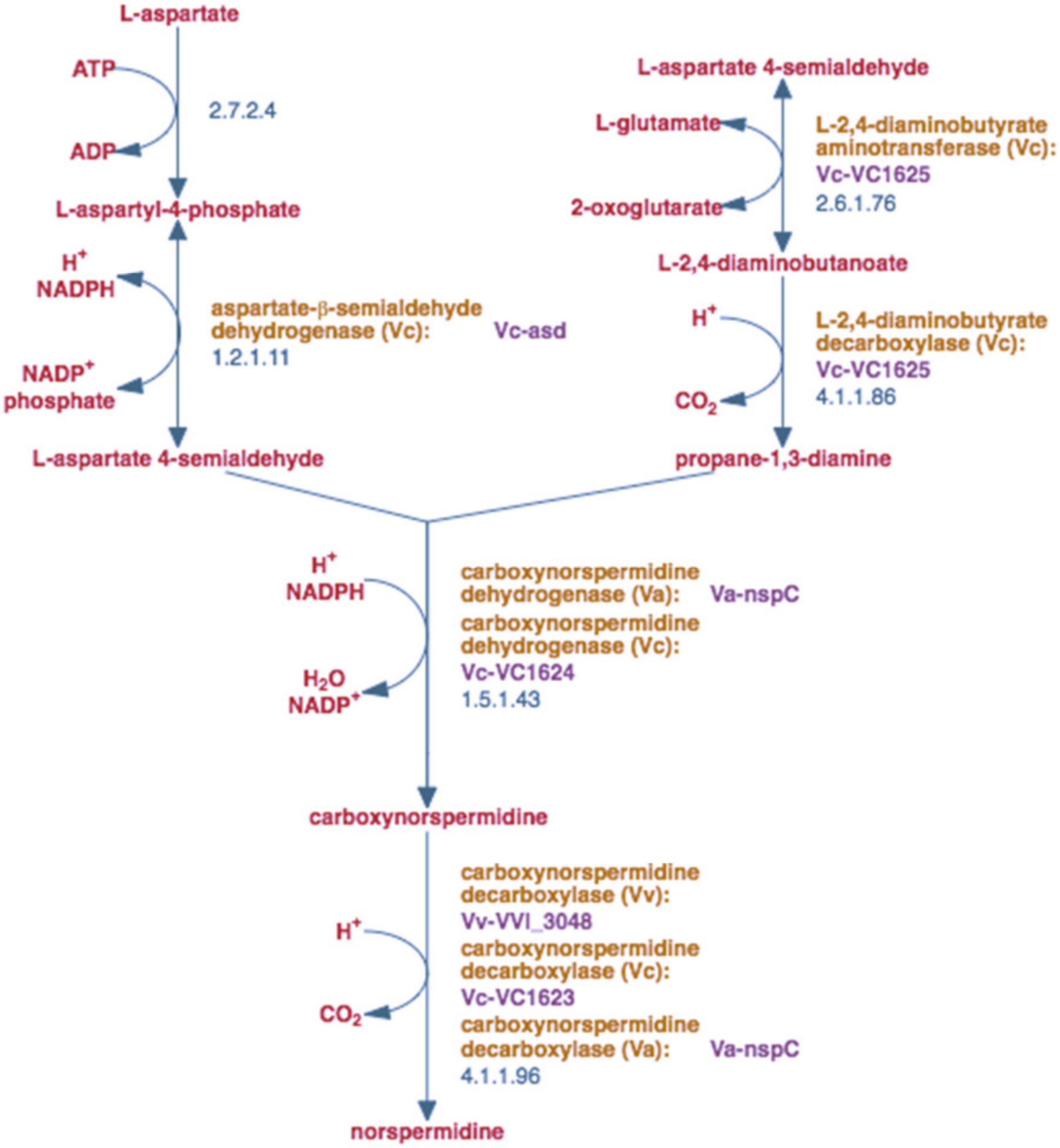

**Supplementary Figure S3:** MetaCyc **Pathway: norspermidine biosynthesis**

(https://biocyc.org/META/NEW-IMAGE?type=PATHWAY&object=PWY-6562&detail-level=2)
